# Universities and COVID-19 Growth at the Start of the 2020 Academic Year

**DOI:** 10.1101/2020.11.25.20238899

**Authors:** Mike Penuliar, Candice Clark, Debbie Curti, Cathy Hudson, Billy Philips

**Affiliations:** F. Marie Hall Institute for Rural and Community Health, Texas Tech University - Health Sciences Center, Lubbock, Texas

**Keywords:** COVID-19, SIR Model, University Health, College Health, Public Health

## Abstract

The global pandemic of 2020 caused by the novel coronavirus of 2019 (COVID-19) has uprooted the education system of the United States. As American colleges and universities try to resume regular instruction for the 2020-2021 academic year, outbreaks have begun to emerge and university towns across the country are now virus hotspots. The current paper provides two studies. First, the current work investigates how the growth of COVID-19 compares in areas with large universities against those without. Results showed markedly increase case growth in counties with large universities at the start of the fall 2020 semester. Secondly, this work provides a highly accessible and modifiable epidemiological tool known as a susceptible-infected-removed model for educational administrators that will allow users to see the impact of COVID-19 historically and predictively. The results of an exemplar model using a large public research university, Texas Tech University, are discussed.

## Introduction

As the novel coronavirus of 2019 (COVID-19) continues to change day-to-day life around the world, a new struggle has come to light in America. After many states relaxed their social distancing measures throughout late spring and early summer, the question of allowing millions of students to return to school became a more pressing concern as the summer wore on. When America went into quarantine in March 2020, schools across the country sent their students home to finish the year through distance learning, with mixed results based on demographics and implementation.^1^ As early as April 2020, researchers were lamenting the negative effects of school closures on their students, ranging from increased food insecurity to decreased academic performance.^2^

Universities were also feeling financial pressures that could only be solved by having an in-person student body. For many colleges, revenue could not be generated from housing or dining plans if there were no students present on campus.^3^ As the year wore on and the pandemic persisted, many Americans grew tired of waiting. Measures were eased and workplaces reopened in summer 2020. Then the pressure was on to open schools again. When August 2020 arrived, many school districts and universities across America caved in and welcomed their students back for in-person learning. Students started moving into dorm rooms for their fall semester despite the growing number of cases in various regions and the obvious inherent risks of being present in a current or potential outbreak. Researchers saw similar factors in university outbreaks as what was seen previously in outbreaks at meatpacking plants, nursing homes, and prisons where multitudes of peoples work and live closely in one locale. ^4, 5, 6^

As of September 2020, multiple schools have opened for the fall semester, only to close within weeks and move back to online learning due to large outbreaks of COVID-19 cases. For example, the University of North Carolina – Chapel Hill, Michigan State, and Notre Dame have moved their fall 2020 classes online after COVID-19 outbreaks early in their semesters, in order to curb the virus’ spread.^7, 8^ Obstinacy is one theory as the University of Illinois, a highly prepared institution with numerous safeguards, had a significant outbreak that grew from COVID-19 positive students who refused to isolate and continued to host or attend social gatherings.^9^

Further compounding the issue, universities implemented different testing and data storage processes for their students’ return as there was no universally accepted protocol.^10^ In Texas, Texas Tech University offered free drive-thru voluntary testing until the first day of school, with subsequent testing under Student Health Services. At Texas A&M University students underwent mandatory testing, resulting in a remarkably high case rate. The University of Texas at Austin and the University of Houston also tested and updated their websites with new case counts every day. On a wider scope, a survey of 500 colleges and universities found that only 27% of universities performed re-entry testing before the start of the semester, and only 20% of universities have committed to routinely test their populations.^11^

It is clear that universities are facing the consequences of their choice to resume university operations as normal as possible with limited resources and guidance. Students who work for universities have expressed their frustration too. Disgruntled graduate student employees of the University of Michigan have organized a strike in response to the university’s COVID-19 response.^12^ At West Virginia University, punitive actions against party hosts were taken,^13^ but it has yet to be seen if this sets any precedence or provides any behavior modification to the student body in the fight against COVID-19.

It is a difficult time, and the current paper hopes to aid university decision-makers in relation to the COVID-19 pandemic by examining it within a higher education framework at two levels: national and local, and by providing a tool to help guide strategic planning for university administrators. First, this work compares COVID-19 growth at the start of the academic school year in counties with large universities against counties without those types of universities. Next, the focus was narrowed to a single large public research university in the American Southwest, Texas Tech University (TTU). With concerned administrators, parents, and students wavering between learning in-person versus the risk of spreading COVID-19, we have also created a highly usable and modifiable epidemiological model that can predict the transmission rate of schools and colleges that pursue in-person learning. The model created for this study allows users to visualize rates of transmission and infection for all educational settings, from elementary all the way to the university setting. Using the information available in this paper and our modeling tool, schools can decide whether to stay open or move to distance learning based on the rate of students transmitting and becoming infected with COVID-19 historically or in the near future.

### Study 1: Counties with Large Universities

For the first study, we analyzed COVID-19’s growth rate in the United States at the beginning of the fall 2020 academic year using counties with large universities (CWLU) against counties without these types of universities, all other counties (AOC). It was hypothesized that there would be an increase in case growth at the beginning of the school year that would contrast to a decrease in case growth elsewhere, due to the movement of many students going back to university towns to resume their education at reopened universities. The current study defined large universities as those with at least 20,000 students enrolled. This study period analyzed case growth rates across four weeks, August 5, 2020 to September 1, 2020.

### Method

United States county COVID-19 case data was collected from *The New York Times*.^14^ University enrollment data was collected from the Homeland Infrastructure Foundation.^15^ New COVID-19 cases from counties with universities that had at least 20,000 students enrolled during the 2018-2019 academic year, CWLU, were compared against counties that did not contain these types of universities, AOC. The categorizing of categories led to 139 CWLUs and 2,965 AOCs. To compare growth rates between county types, the daily percent change of each county was calculated across four individual weeks: Week 1: August 5, 2020 to August 11, 2020, Week 2: August 12, 2020 to August 18, 2020, Week 3: August 19, 2020 to August 25, 2020, and Week 4, August 26, 2020 to September 1, 2020.

### Results

A 4 (week) × 2 (county) repeated measures ANOVA was performed using county type as the between-subjects factor and daily percent change as the dependent variable. Results showed a significant interaction, *F*(2.83, 8765.94) = 3.07, *p* = .03, *η*_*p*_^2^ = .001. See Table 1 for *M, SD* of the average daily percent change in COVID-19 cases across weeks.

**Table 1.** Means and Standard Deviation as a function of Week and County Type

| **Week** | **County Type** | **M** | **SD** |
| --- | --- | --- | --- |
| Week 1 | CWLU | 0.0116 | 0.0070 |
|  | AOC | 0.0202 | 0.0292 |
| Week 2 | CWLU | 0.0098 | 0.0063 |
|  | AOC | 0.0169 | 0.0298 |
| Week 3 | CWLU | 0.0113 | 0.0124 |
|  | AOC | 0.0152 | 0.0222 |
| Week 4 | CWLU | 0.0138 | 0.0161 |
|  | AOC | 0.0144 | 0.0236 |

Figure 1 shows the changes across time between county types. From Weeks 1 to 2, both groups showed decreasing growth in new cases. This changed in Weeks 2 to 4, where there was an increasing trend of new cases in CWLU compared to AOC, indicating that the influx of students to these types of counties potentially caused a surge of new cases. In comparison, AOC continued to have decreased growth in new cases.

**Figure 1.**
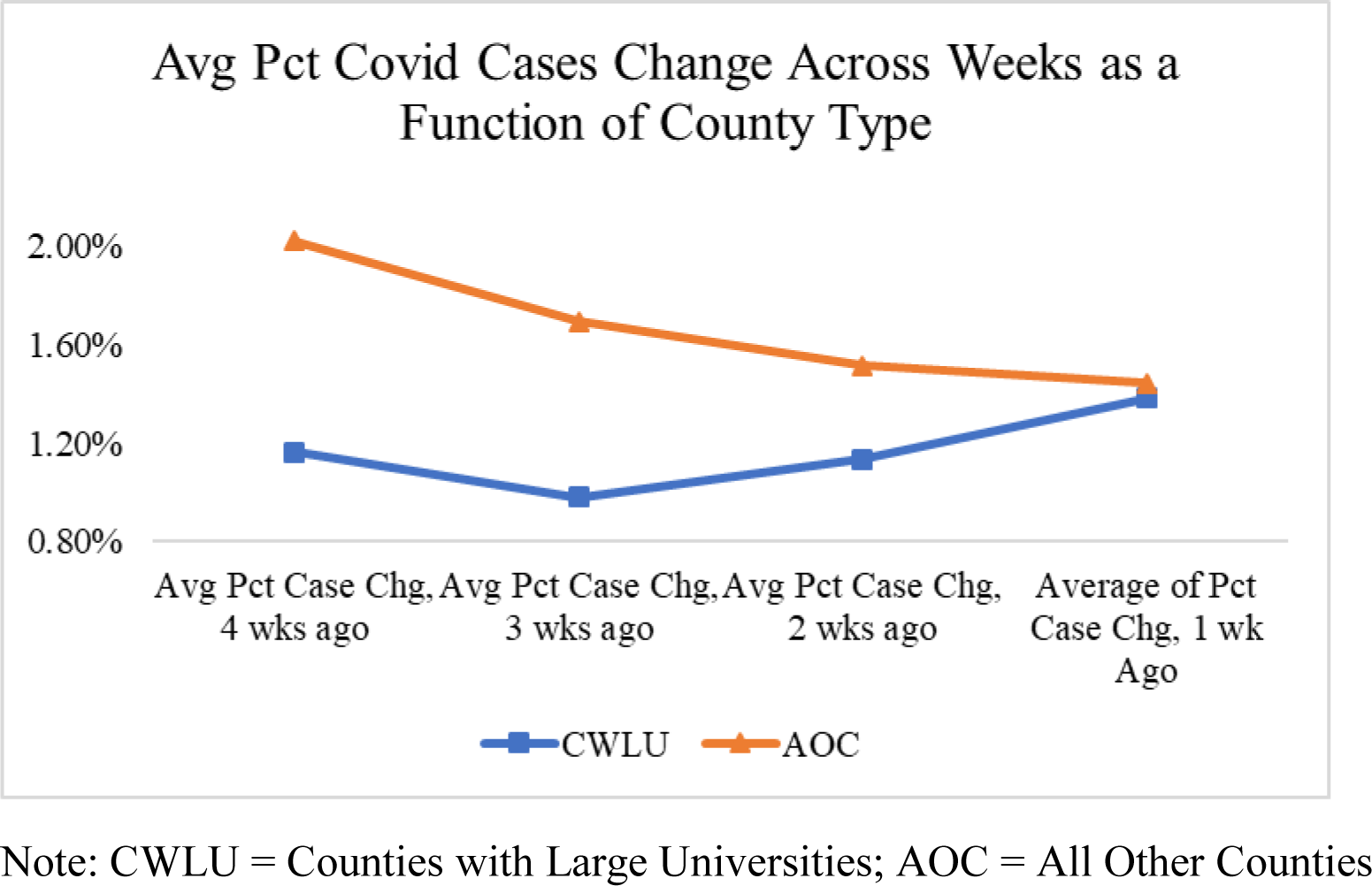
Average Percent Covid Cases Change Across Weeks as a function of County Type

### Discussion

After segmenting counties based on the criteria of the presence of a large university, a distressing trend for the start of the 2020 academic year started to emerge. University towns across the United States appeared to be experiencing accelerated case growth. These trends are not surprising as college students from around the nation returned to university campuses. Analogous to previous hotspots of prisons and meatpacking plants, universities have many people interacting in enclosed, crowded, or cramped spaces, which may contribute to increased case growth. Many students are also experiencing their first brushes with being highly autonomous and away from home which allows them the opportunity to engage in behaviors some may regard as risky to their health.

Financial pressures from administrators are likely one key reason universities decided to resume in-class learning.^16^ A university is very much a business, and digital instruction and remote courses do not provide all the revenue universities need to function. Dorm rooms, dining, bookstores, and athletic activities are missed income if universities continue to teach primarily online. Without the funds generated from a traditional college experience, paying educators, administrators and staff may be difficult.

These are some of the reasons why universities decided to open, and as students returned and started their college classes, *The New York Times* published an article in early September 2020 containing two lists: top metro areas experiencing the biggest outbreaks at the moment and metro areas that would be experiencing big outbreaks in the near future.^17^ Of the metro areas lists, nine out of ten had colleges or universities with greater than 20,000 students enrolled. The following study narrows the focus to one university to track the outbreak at the start of the fall 2020 academic year. Lubbock, Texas, home to Texas Tech University, was present on both lists.

### Study 2 – University Susceptible-Infected-Removed Model

Study 2 narrowed the scope of Study 1 and solely focused on a Susceptible-Infected-Removed (SIR) model for TTU, a large public research university in West Texas. This study was conducted to further understand the pandemic at a local level and within a higher education framework. SIR models are relatively simple epidemiological tools that can be easy to create. These models divide people into compartments with the simplest models only having three groups: susceptible, infected, and removed (sometimes called recovered). The current study advocates for building an SIR model within spreadsheet software such as Microsoft Excel, to track and forecast potential cases for an educational institution of interest.

The particular model created for this study grants users the ability to forecast potential scenarios (e.g., increased or decreased social distancing) for an educational intuition using two important factors: the infection rate (β) and the recovery rate (γ), and dividing the former by the latter yields the basic reproductive number, R_0_. An R_0_ > (greater than) 1 is evident of a growing pandemic as each infected person spreads the disease to more than one person on average. The opposite is true for R0 < (less than) 1 as each person will infect less than one person on average. Both β and γ can be estimated from analyzing local trends or hypothesized for future events. Analogous to true to life scenarios, infection rates can be altered in the study’s model as per the user’s observations or hypotheses. For the current study, TTU was used to showcase the usability and ease of this model for educational settings. Though this particular higher education setting was selected, users of this model may modify the parameters to fit any education setting, district, or level they desire (e.g., elementary, middle, or high school).

### Method

### Data

The TTU SIR Model in this study was created using public data from the TTU COVID-19 and Lubbock COVID-19 dashboards collected daily from August 23, 2020 to September 10, 2020.^18, 19^ There was no institutional review board (IRB) review as this was all public data. University population estimates were collected from Texas Tech University.^20, 21^ Community population estimates and COVID-19 cases to estimate infection rates were collected from the U.S. Census and the Lubbock COVID-19 Dashboard, respectively.^22^ School holidays and events were also notated within the spreadsheet to help ascertain possible surges. Finally, the estimated percentage for non-infectious infected, asymptomatic infectious, and symptomatic infectious were culled from the CDC in August 2020 and used in various components in the model.^23^

### SIR Model

The SIR model by Penuliar et. al. was modified for this study.^24^ Appendix A provides a truncated view of the model’s spreadsheet. The full file can be downloaded at github.com/mpenuliar-ttuhsc/SIR, where users can see the formulas in a spreadsheet setting, alter it to fit their research interests, forecast, and assess model fit. The compartment formulas and overall process for the current study’s SIR model can be found in Table 2 and Figure 2, respectively. Subscript n refers to the current time step and subscript n-1 refers to the previous time step. The sum of S, I, and R at each time step was always equal to the study population.

**Table 2.** Susceptible-Infected-Removed Formulas

| Variable | Abbreviation | Formula |
| --- | --- | --- |
| Susceptible | $S_n$ | $S_{n-1} - I - S_n - I - H_n$ |
| Infected from School | $IS_n$ | $(S_{n-1}/S) * (\beta * I - I_{n-1})$ |
| Infected from Home | $IH_n$ | $(S_{n-1}/S) * (\beta_c * I_c)$ |
| Total New Infected | $TNI_n$ | $IS_n + IH_n$ |
| Newly Infectious Infected | $NII_n$ | $TNI_n * .9$ |
| Newly Non-Infectious Infected | $NNII_n$ | $TNI_n * .1$ |
| Newly Asymptomatic Infectious | $NAI_n$ | $TNI_n * .3$ |
| Newly Symptomatic Infectious | $NSI_n$ | $TNI_n * .6$ |
| Infectious Infected | $II_n$ | $II_{n-1} + (NII_n) - (II_{n-1} * \gamma)$ |
| Removed | $R_n$ | $R_{n-1} + (I_{n-1} * \gamma)$ |

**Figure 2.**
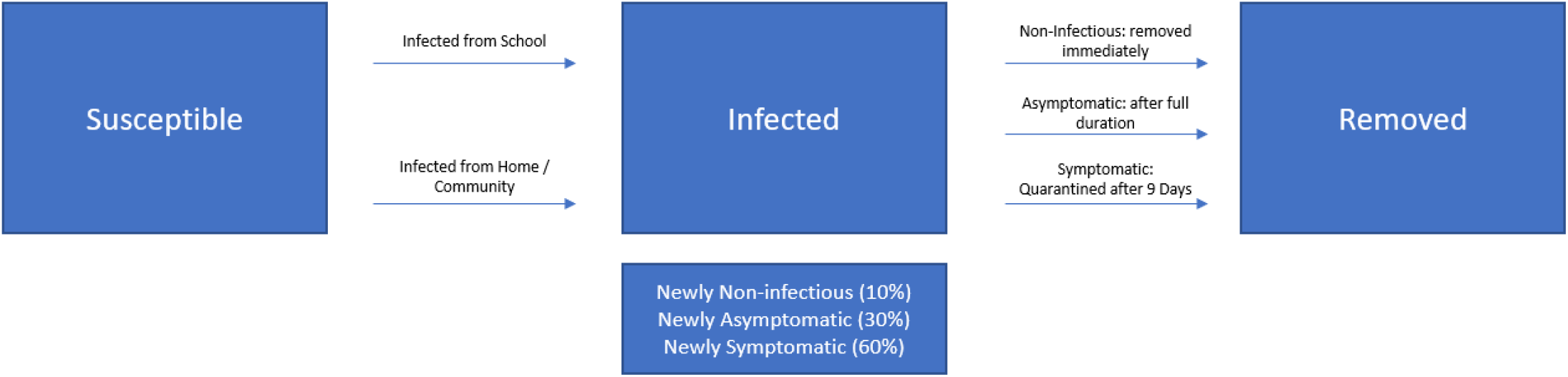
Susceptible – Infected – Removed Flowchart

Though the current model is based on a basic SIR framework, there were special considerations for infectious (i.e., non-infectious and infectious) and symptomatic (i.e., symptomatic and asymptomatic) types. These categories are Newly Non-Infectious Infected (NNII), Newly Asymptomatic Infectious (NAI), and Newly Symptomatic Infectious (NSI). The three categories summed to Total New Infected (TNI) at each time step. Based on the August 2020 CDC estimates mentioned previously, NNII is 10% of TNI, NAI is 30% of TNI, and NSI is 60% of TNI.

Also unique to this model is a quarantine implemented for symptomatic infections on the 8th day of infectiousness, which transferred them from the infectious category and into the removed category. This day was chosen as it is the mean amount of days for symptom appearance.^25^ In comparison, asymptomatic carriers remained in the infectious category for the duration of their disease. Initial infectious infected was equal to 90% of the starting infected number and the remaining 10% were added to the initial removed category.

The current study’s SIR model started with 53 infected and 50 recovered within the given population, as this was the case count the day before the start of the fall 2020 term.^26^ The starting infection and recovery parameters within the model were β_1_ = .13, γ =.045, and β/γ = R_0_ = 2.89, meaning that each infected person at the university is presumed to have infected 2.89 more people at the start of the model. Infections could also be acquired from interactions with a close relation from the community (e.g., a close non-university affiliated social connection, family member, or a non-university affiliated roommate), β_2_ = .70, which is higher than β_1_ due to the proximity and nature of these types of interactions. In the model, the percent active in the community at any given time was set to .48%. This was based on the percentage of active cases in Lubbock County prior to the start of the fall semester. Initial starting parameters were derived from reviewing the historical data of both the university and Lubbock County across time and assessing model fit when using estimated infection rates. The SIR model building process established a model fit benchmark of at least *R*^2^ = .90 between historical cases and predicted cases for each SIR compartment. All starting metrics can be seen in Table 3 and in the full model file.

**Table 3.** TTU SIR Starting Parameters

| **Metric** | **Starting Number** |
| --- | --- |
| SIR Total | 28,310.7 |
| ($\beta_1$) - S infected/day - School | 0.130 |
| ($\gamma$) - Recovered/day | 0.045 |
| $R_0$ | 2.89 |
| Roommate or close social contact per day | 0.25 |
| Total Close Social Contact per Susceptible | 7077.7 |
| Confirmed Active for Model | 1475 |
| Total County Cases on 8/24/20 | 6918 |
| Total County Recovered on 8/24/20 | 5349 |
| County Dead on 8/24/20 | 94 |
| Remaining County Susceptible on 8/24/20 | 303,651 |
| Pct active community infected for model | 0.48% |
| County sans TTU Students | 275,340.3 |
| Close Contacts Infected at start of model | 34.21 |
| Chance of Roommates or Close Contacts Infecting Student | 0.7 |
| Total Students and Staff | 40,591 |
| Percent S of Total Pop | 70% |

The SIR modeling tool in this study allowed users to increase or decrease infection rates based on historical or hypothesized events. For example, the TTU model provided two hypothesized future scenarios that modified the infection rate for set periods of time. These conditions were akin to best-case and worst-case scenarios. The model ends on the last day of semester, December 9, 2020.

### Results

#### Historical Cases

To best model future scenarios, a strong fit between historical and predicted cases within the SIR model was necessary. First, the total confirmed cases were regressed on the TTU SIR model’s total predicted cases, *R*^2^ = .99, *F*(1, 17) = 1179.90, *p* < .001, *RMSE* = 39.78, and the analysis found a strong model fit (Figure 3). Next, a regression analysis was performed on the predicted versus observed active case data, *R*^2^ = .94, *F*(1, 17) = 265.09, *p* < .001, *RMSE* = 114.59, and the analysis found good fit as well (Figure 4). Finally, the actual recovered cases were analyzed against the predicted recovered (Figure 5). The final regression analysis showed good model fit, *R*^2^ = .99, *F*(1, 17) = 1136.35, *p* < .001, *RMSE* = 107.82. Taken together, these three linear regression analyses indicated strong model fit and that the starting model parameters led to an accurate estimation of the COVID-19 reality at TTU. Importantly, after nearly three weeks from August 24, 2020, the start of the 2020 school year, it is clear that there was an increasing trend of confirmed infected COVID-19 cases at TTU which agrees with the expectation that the returning of students and employees at the beginning of August 2020 would lead to rapid case growth.

**Figure 3.**
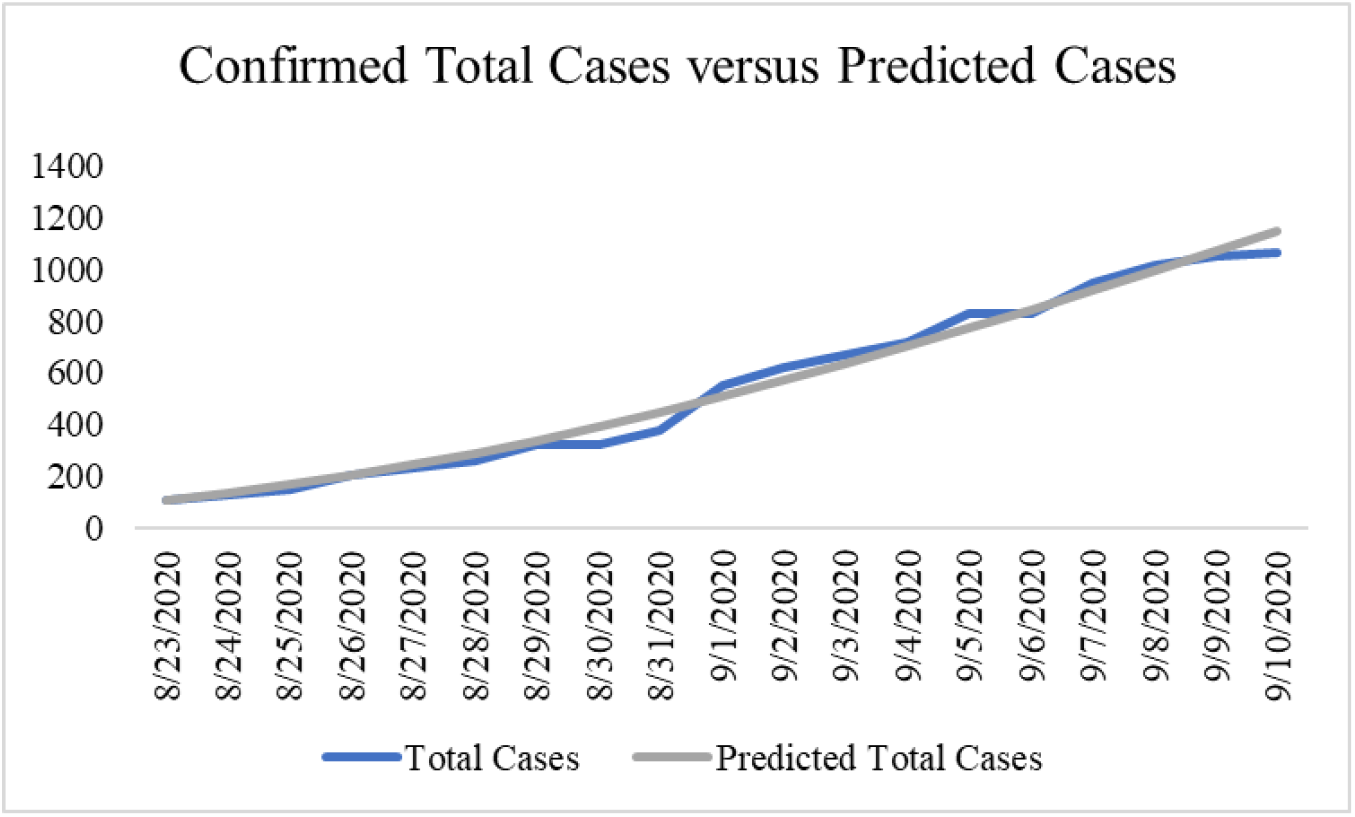
Confirmed Total Cases Plotted Against Predicted Total Cases

**Figure 4.**
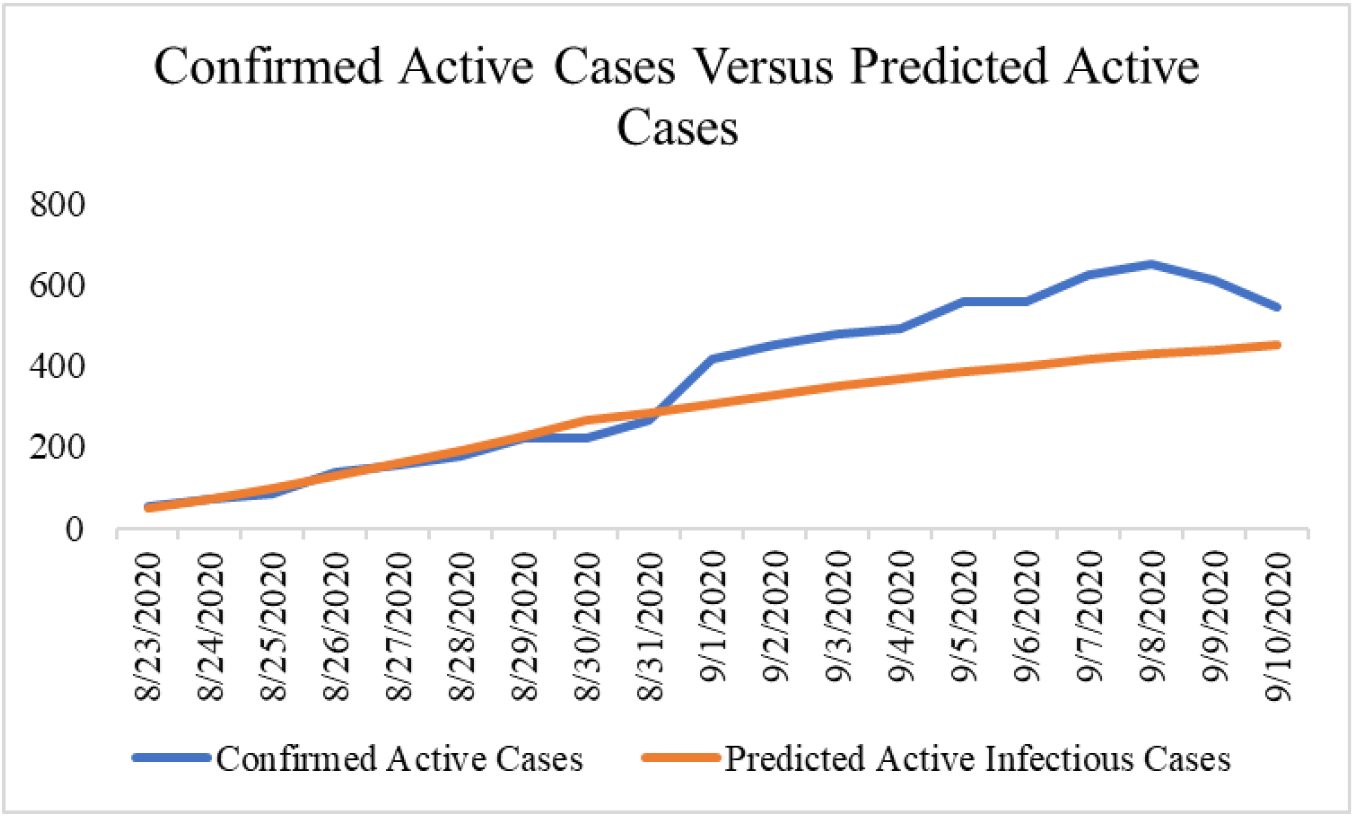
Confirmed Active Cases Plotted Against Predicted Active Cases

**Figure 5.**
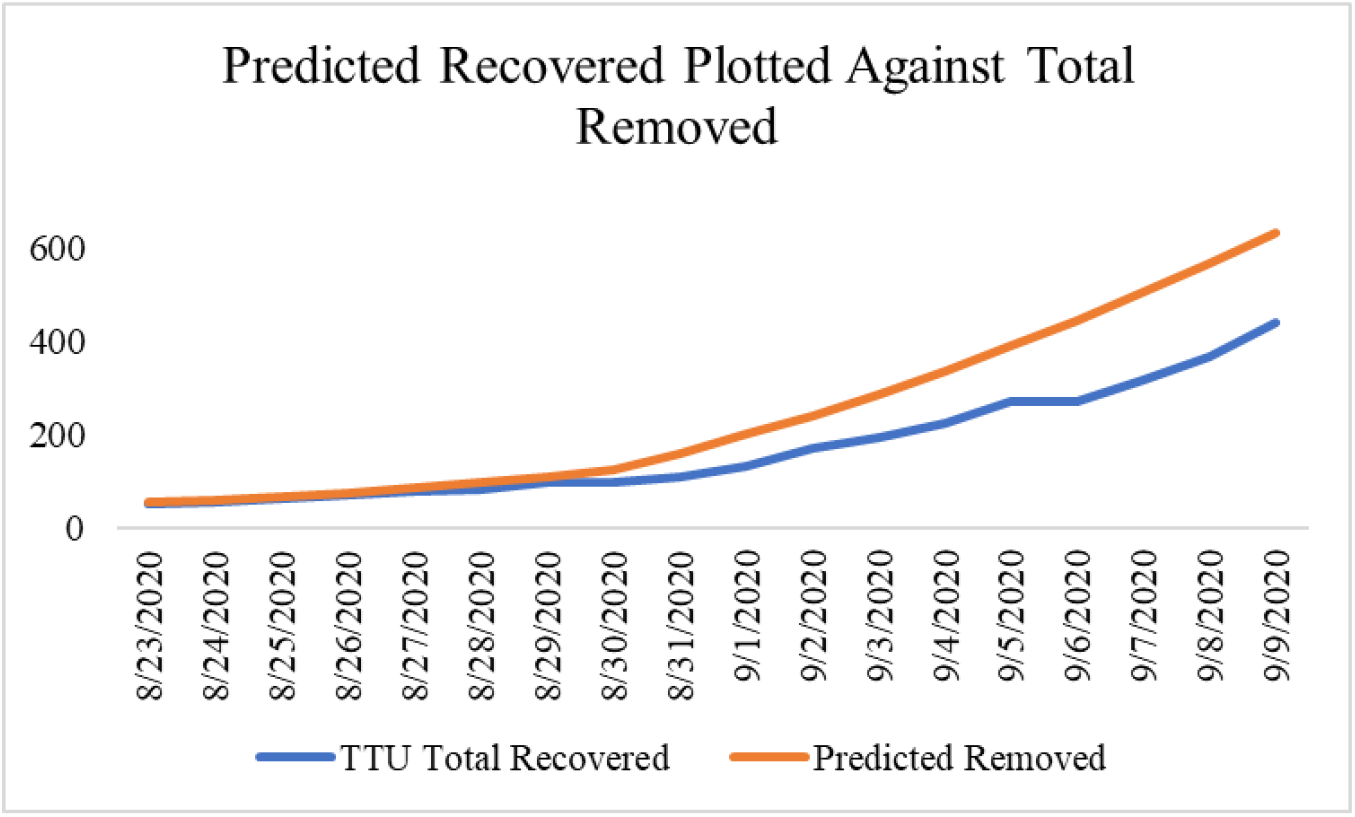
Predicted Removed Plotted Against Total Recovered

#### Future Cases

Fortunately, the model’s predicted infections to recoveries ratio (*new infection/new recoveries*) reached below one on September 1, 2020, indicating that predicted recoveries outpaced the predicted new infected from that point forward. This likely denotes that there would be a peak in active infections a few weeks into the Fall 2020 semester, and that peak would decline as the semester wore on. Another promising statistic was the total case doubling time (*ln*(*2*)/(*slope*(*ln*(*previous 7 day cases*)))) (Figure 6). In most instances, the doubling time was less than one week but has grown quickly after September 7, 2020. On September 10, 2020 the doubling time was 10 days. Finally, Figure 7 plots the three-day change average of log total cases. This plot shows a strong downward trend which signifies that case growth may be decelerating. Taken together, these statistics signified promising news that a slowing of new cases may be approaching.

**Figure 6.**
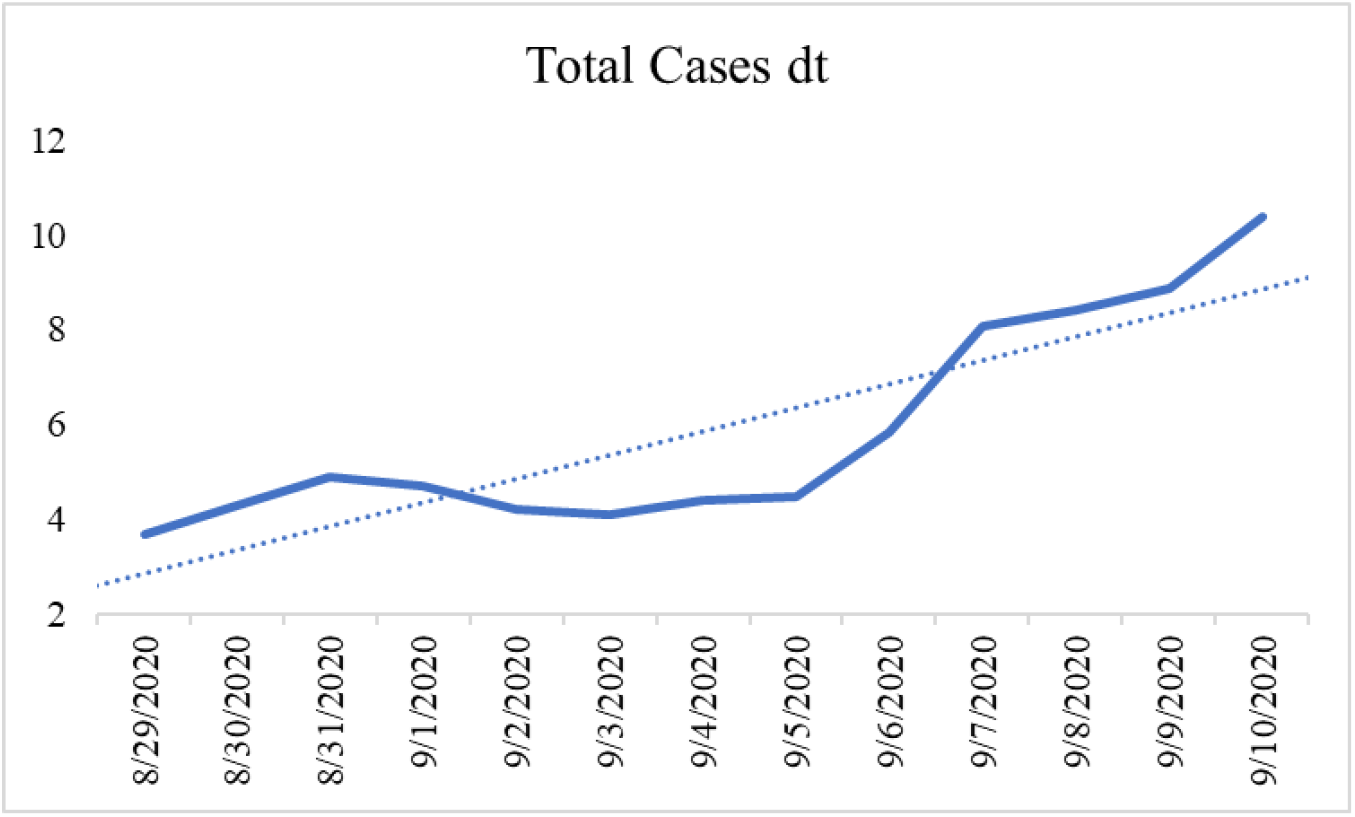
Total Case Doubling Time – TTU SIR Model

**Figure 7.**
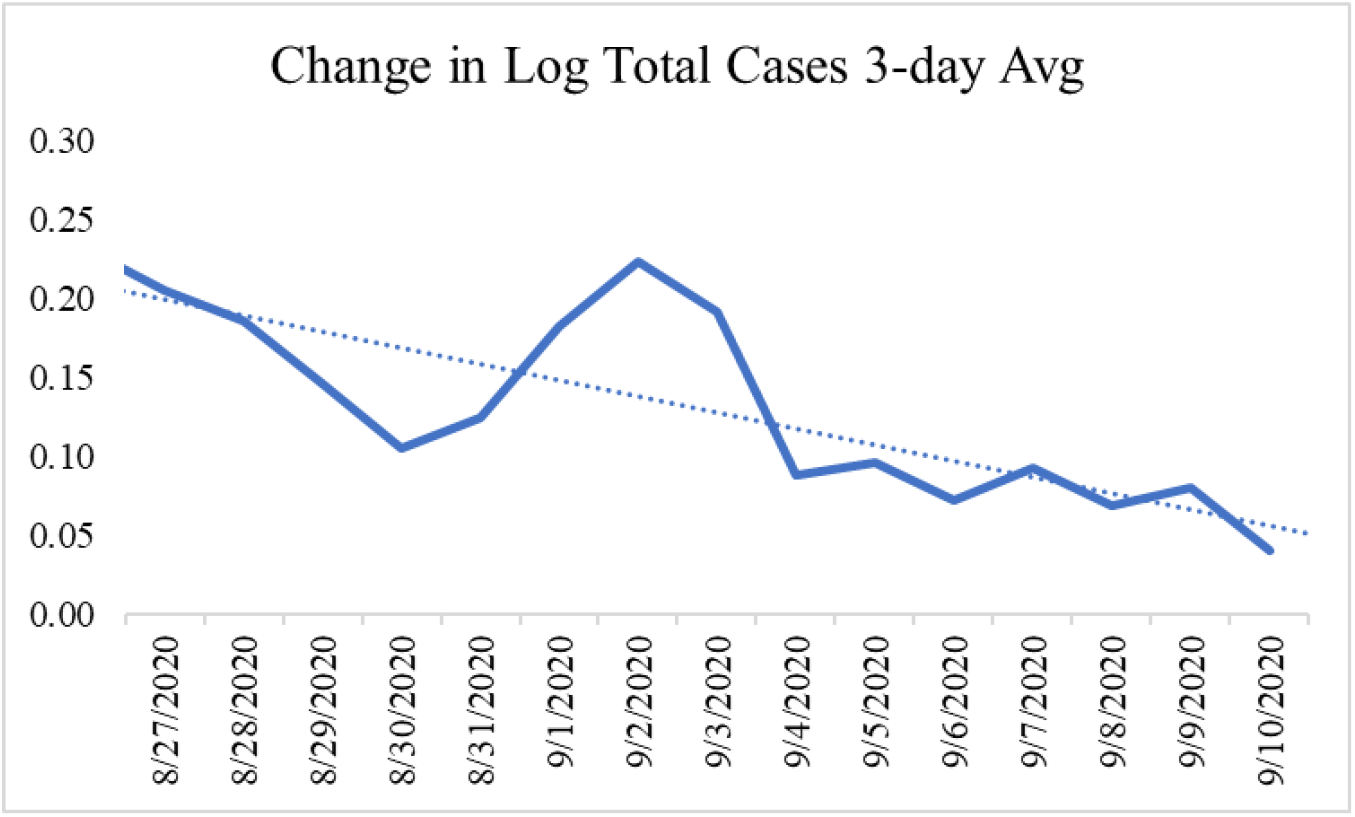
Change in Log Total Cases – 3 Day Average – TTU SIR Model

#### Potential Scenarios

Figure 8 shows the predicted infected cases for the 2020 fall academic semester, August 23, 2020 to December 9, 2020 across two scenarios: no mitigation and successful mitigation. These conditions were based on updated infection rates starting on September 11, 2020, a three-day lag after Labor Day weekend. Table 4 lists key events and the changes in infection rate across the conditions.

**Table 4.** Key Events, Dates, and Infection Rates Across Groups

| Key Dates | Event | No Mitigation $\beta$ | Mitigation $\beta$ |
| --- | --- | --- | --- |
| 8/23/2020 | First Day of Fall 2020 Semester | .130 | .130 |
| 9/11/2020 | Post Labor Day Weekend | .163 | .034 |
| 9/12/2020 | First Football Game | .163 | .034 |
| 10/01/2020 | End of Post Labor Day Spike (No Mitigation Condition) | .130 | .034 |
| 10/30/2020 | Halloween Events | .163 | .034 |
| 11/20/2020 | Thanksgiving Holiday (No Mitigation Condition) | .034 | .034 |
| 11/30/2020 | Return from Thanksgiving of Fall 2020 Semester | .163 | .034 |
| 12/09/2020 | End of Fall 2020 Semester | .163 | .034 |

**Figure 8.**
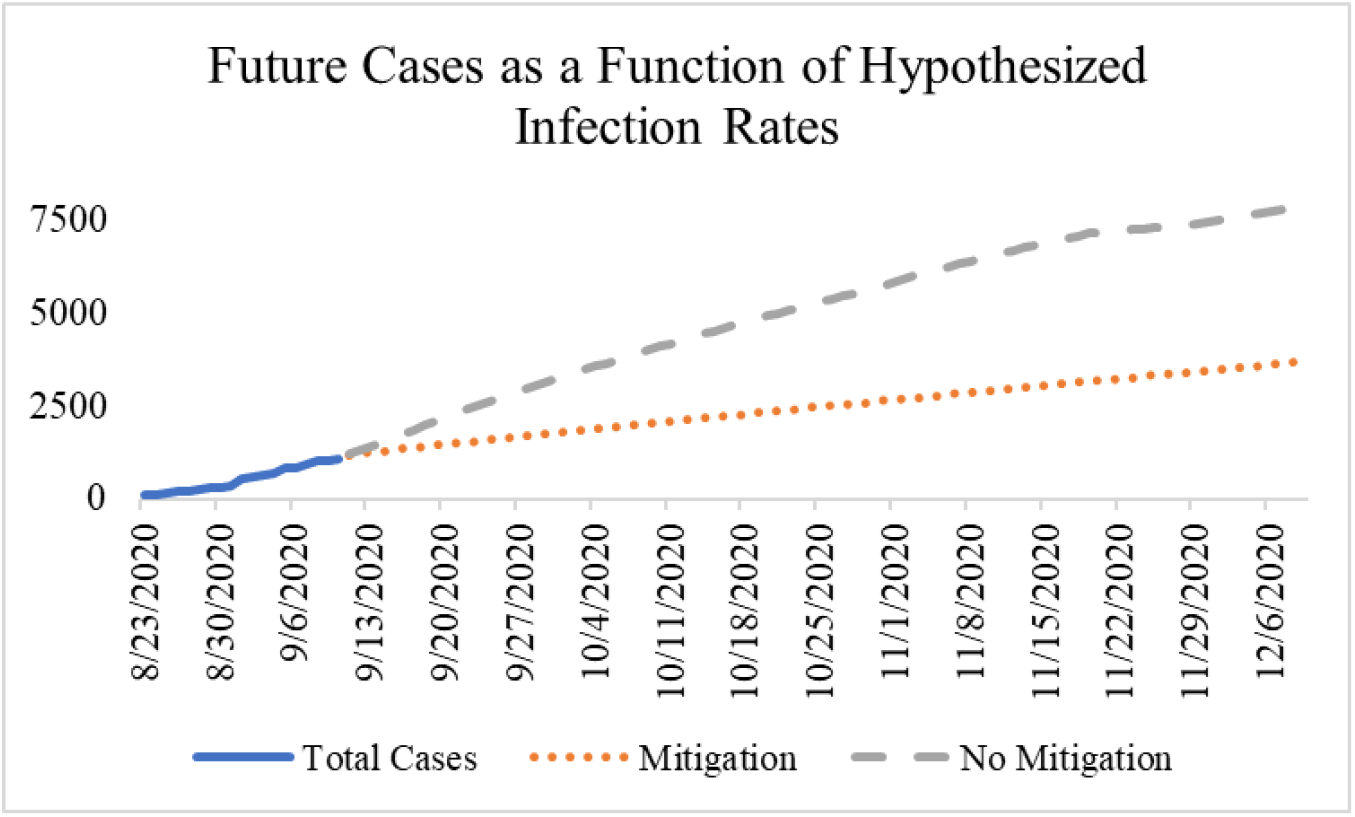
Potential Scenarios - TTU SIR Model

The no mitigation condition was characterized by three infection spikes, β_no_mit_ = .163, R_0_ = 2.89, during the fall 2020 term. The first spike started on September 11, 2020 and lasted for 10 days, overlapping with TTU’s first in-person football game before returning to β_1_ = .13. The second spike started on October, 30, 2020, aligning with Halloween celebrations, and lasted for five days. The infection rate returned to the original rate, β_1_ = .13, until the Thanksgiving break, β_break_ = .034, which provided a reduced rate that lasted for 10 days. On November 30, 2020 when the students returned, with many potentially transporting the virus, the rated returned to β_no_mit_ = .163, which lasted until the end of the semester. In the no mitigation condition, active infectious cases would peak at 698 on September 21, 2020, and the total cases by the end of the Fall 2020 term would be 7,982.

The successful mitigation condition, β_mitigation_ = .034, R_0_ =.75, started on September 11, 2020 and lasted until the end of the term. If the increase in total case doubling time continues, this trend will possibly emerge. In the successful mitigation condition, the model’s peak infectious cases was 451 on September 10, 2020. The total predicted cases by the end of the fall 2020 semester was 3,709. This condition will possibly appear if enhanced social distancing and hygiene behaviors in the university’s population were routinely performed in response to the initial exponential growth in cases at the beginning of the semester.

### Discussion

At the start of the 2020 academic year, COVID-19 cases increased dramatically at TTU. This was to be expected as students returned to the university to resume their education and employees of the university returned to help facilitate that education. Though safeguards such as enhanced sanitation, health screenings, access to personal protective equipment, and social distancing guidelines were put in place throughout the university in preparation for the return of students, cases still climbed. ^27, 28^

One factor contributing to this climb may be the nature of the university environment that fosters collaborations, connections, and socialization, which are key in the traditional university experience. Unfortunately, as people connect, the chance of them spreading the virus increases. Though controls to slow the spread exist within the university, enforcement of those controls may not be uniform or adequate across departments. Next, once students leave campus it is their choice to do as they wish with their free time. If that free time consists of socialization at a large gathering without proper face coverings or hygiene behaviors, there is a chance that someone may unwittingly spread the virus. Additionally, there may be mixed messages from the university and what students see around the community or in local businesses which may not inspire confidence among a student population to follow public health best practices.^29^ Finally, the laissez-faire attitude of some young people may be due to the common perception of this demographic as being generally hardier compared to those with pre-existing conditions or advanced age. Though TTU is home to many young adults, those teaching these students and working at the university as well as the residents of the city of Lubbock vary in age and health condition, and any COVID-19 spikes in the campus will ultimately affect the faculty, staff, and the surrounding community in some way.

The presented modeling tool with the inputted parameters has shown excellent fit and the danger of not mitigating the spread of COVID-19. It is important to understand that even at full mitigation efforts, the spread of the virus can still occur and proper precautions must be adhered. Fortunately, though cases have spiked early in the academic year, there are some promising signs in the data. Specifically, the increased doubling time and the decreased growth of the log total COVID-19 cases. This could indicate the rapid growth phase of the initial return to campus may be approaching an end. Regardless, it is still imperative for students and staff to continue to mitigate the growth of new cases as best as possible. A mitigated growth curve carried out by excellent hygiene behaviors, social distancing, face coverings, and proper quarantining is preferable to an unmitigated curve that could lead to deleterious results.

### Limitations & Future Directions

Taking the evidence from the SIR modeling tool and cases from universities around the nation, there appears to be a strong connection between the return of students to universities at the start of the 2020 academic year and an increase in case growth. The results of the two studies here are quasi-experimental and correlational in nature, full causality cannot be determined without a true experimental design. This opens avenues for future research on how to fully stop the spread of COVID-19. Some studies have already contributed to this effort. Specifically, the efficacy and types of masks to beat this pandemic.^30^

The SIR modeling tool created for this paper was tailored for a university setting. Future researchers may use this tool for other education levels such as elementary or high school. Important to note, this tool relies on historical case data to assist in forecasting future cases. If a school does not publish their case data, the estimates gathered from this tool may be limited in application. Future researchers interested in adjusting the TTU SIR model parameters are able to do so as the modeling tool has high accessibility. For instance, if a researcher believes that there will be a larger infection spike in relation to Halloween, October 31, 2020, they can easily modify the model and view those results. The model also uses a fixed active infection rate for the community. Future researchers may wish to make this a dynamic rate in future iterations of this modeling tool.

## Conclusion

The desire to return to pre-pandemic normality is understandable. The year 2020 has not been kind to humanity. Not only did this year bring a pandemic but numerous tragic and disconcerting events that have shaken the core of many people living in the United States. In the true vein of the land of the free and home of the brave, students and non-students alike are out and about across many towns trying to survive as they search for a glimmer, a sign that everything will be okay. They are at shops, restaurants, events, malls, and a variety of locations trying to bring some aspect of joy and normalcy to their lives or their family’s lives. Other young people may not be partying, but simply working in essential or service jobs to satiate the basic needs of food and shelter for their loved ones.^31^ These roles, often filled with young people, have been integral in keeping the economy from shuddering to a complete stop. Research has also indicated that young adults reported more mental strain in response to the pandemic compared to older demographics as they are especially worried about potential financial losses, job productivity, and the health and safety of loved ones.^32^

The unfortunate truth for all is that we are living through a pandemic, and the virus on our shores does not care about race, age, sex, politics, or the belief of its existence. COVID-19’s primary goal is to replicate and spread, and it uses the human body as a host in order to achieve that goal. One of the easiest ways to spread is being present in large gatherings of unfamiliar people, especially without the proper use of face coverings.

It is understandable why some university students may downplay the threat of the virus. Young adults may get sick, but many in this age bracket will not die from COVID-19 as co-morbid health conditions are not as common within this demographic, relative to other groups. Regrettably, as the virus spreads through young adults it will likely trickle down to the surrounding university and community. One of those infections may find its way into a body that is less adept at fighting it due to their unique biology or personal circumstance.

Teachers, staff, family members, and community members come in a variety of ages and health conditions. Adding to the over 200,000 COVID-19 deaths at the time of writing would only further escalate that tragedy. Every single death as a result of the pandemic has devastated a family in some way. As we continue to reopen our institutes of learning it is important to contribute our part to protect ourselves, our loved ones, our students, and our fellow neighbors which will in turn keep the community healthy and the schools open.

Disclosure Statement – No financial interests or benefits have arisen from this research or the application of it.

Funding – This project was unfunded.

Model File – The SIR Model Template is available on github – www.github.com/mpenuliar-ttuhsc/SIR.

## Supporting information

SIR - School Modeling Tool

## Data Availability

United States county COVID-19 case data was collected from The New York Times. University enrollment data was collected from the Homeland Infrastructure Foundation. University population estimates were collected from Texas Tech University (TTU). Community population estimates and COVID-19 cases were collected from the U.S. Census and the Lubbock COVID-19 Dashboard, respectively. COVID-19 TTU dashboards provided the university's case numbers.

https://github.com/nytimes/covid-19-data

https://hifld-geoplatform.opendata.arcgis.com/datasets/colleges-and-universities-campuses

https://www.depts.ttu.edu/communications/emergency/coronavirus/

https://ci.lubbock.tx.us/departments/health-department/about-us/coronavirus-disease-2019-covid-19

https://www.cdc.gov/coronavirus/2019-ncov/hcp/planning-scenarios.html

http://www.texastech.edu/stories/19-11-record-enrollment.php

https://www.census.gov/quickfacts/lubbockcountytexas

https://ttucovid19.com/#/total

## Appendix A

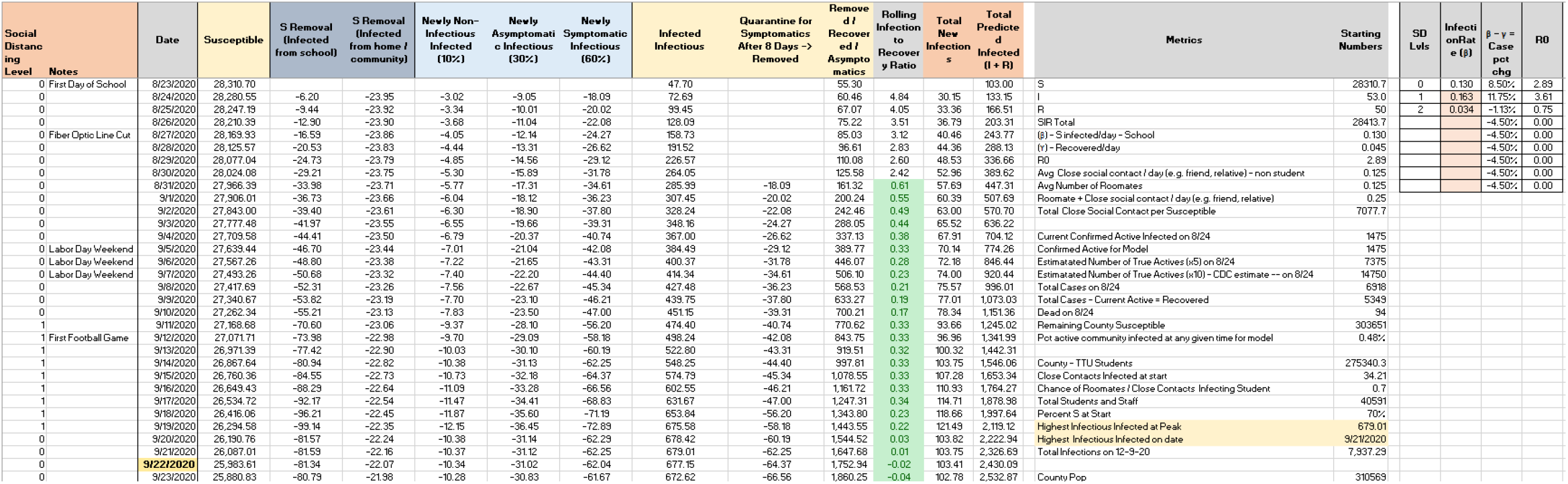

